# Evaluation of the Efficacy and Safety of Combination Therapy of Vamha and Myrha in the Management of PMOS: An Open-Label, Randomized, Multicentre, Comparative, Prospective Clinical Study

**DOI:** 10.64898/2026.08.20.26360875

**Authors:** Aarati Patil, Rahul Barathe, Deepali Murlidhar Tate, Keerti Kate, Shishir Pande, Nitin Gawande, Amruta More, Swapnali Mahadik, Ritika Singhvi, Kshitija Berde

## Abstract

**Introduction:** Polyendocrine metabolic ovarian syndrome (PMOS), formerly known as polycystic ovary syndrome (PCOS), is a common endocrine disorder affecting women of reproductive age. Besides reproductive and metabolic disturbances, PMOS negatively impacts psychological well-being and quality of life. Despite available treatment options, there remains a need for safe and effective therapies that improve both clinical symptoms and fertility outcomes.

**Aim:** To compare the efficacy of VAMHA and MYRHA tablet combination therapy with standard non-hormonal therapy in restoring regular menstruation. Secondary objectives included assessment of ovulation, menstrual symptoms, polycystic ovarian morphology, hormonal and metabolic parameters, anthropometric measures, and skin manifestations.

**Study Design:** Open-label, randomized, multicentre, prospective comparative clinical study.

**Methods:** Seventy-one women with PMOS were randomized to Group A (n=37) or Group B (n=34). Group A received VAMHA and MYRHA tablets (2 tablets each), while Group B received Metformin 500 mg plus Myoinositol 600 mg (1 tablet), twice daily for 180 days. Data were recorded in Case Report Forms.

**Statistical Analysis:** Continuous variables were summarized using mean and standard deviation, while categorical variables were expressed as frequencies and percentages. Appropriate statistical tests, including Chi-square, were used. A p-value ≤0.05 was considered significant.

**Results:** Significantly more participants in Group A achieved regular menstrual cycles than Group B (31 vs. 22; p<0.05). Ovulation occurred in 16 participants in Group A compared with 6 in Group B (p<0.05). Both groups showed significant improvement in menstrual irregularity and related symptoms. Significant reductions in Anti-Mullerian Hormone (AMH), fasting insulin, and body mass index (BMI) were observed in both groups (p<0.05). Resolution of polycystic ovarian morphology occurred in 13 participants (38.23%) in Group A and 10 (33.33%) in Group B. Both treatments were well tolerated with no major safety concerns.

**Conclusions:** VAMHA and MYRHA combination therapy was superior to standard non-hormonal therapy in improving menstrual regularity and ovulation. It also produced favourable metabolic, hormonal, and ultrasonographic outcomes, suggesting its potential as a safe and effective option for comprehensive PMOS management and fertility enhancement.

## 1. Introduction

Polyendocrine metabolic ovarian syndrome (PMOS), previously termed as polycystic ovary syndrome (PCOS), is a common endocrine disorder that affects women of childbearing age. It is typically characterized by chronic anovulation, hyperandrogenism (HA), polycystic ovarian morphology and irregular menstrual cycles (IMC), including oligomenorrhea or amenorrhea [1]. The exact etiology of PMOS remains unclear; however, it is believed to result from a complex interplay of genetic and environmental factors. Women with PMOS are at an increased risk of developing several comorbidities such as insulin resistance, type 2 diabetes mellitus, dyslipidemia, hypertension, infertility, endometrial cancer, and other metabolic disorders affecting mental health outcomes and reducing quality of life [2–4]. The global burden of PMOS is increasing at a high rate because of a sedentary lifestyle, increasing pollution and high intake of junk food [5,6]. In 2021, more than 65 million women were affected by PMOS globally, nearly double earlier estimates, representing an approximately 89% increase over the past three decades [7]. The reported prevalence within India varies widely depending on the population and diagnostic criteria used, ranging from around 3.7% up to 22% in different studies and a pooled prevalence of approximately 10% based on Rotterdam criteria.

The main principle of current conventional treatment is to manage the symptoms and avoid long-term morbidity [8]. These conventional measures primarily include lifestyle modification, insulin-sensitizing agents such as metformin, combined oral contraceptive pills (COCPs), and ovulation-inducing agents. While these therapies are effective in managing specific symptoms, they often target only selected aspects of the syndrome and may be associated with adverse effects, contraindications, poor long-term adherence, or limited efficacy in some participants. Metformin is commonly associated with gastrointestinal intolerance, whereas COCPs may not be suitable for women seeking conception and may increase the risk of certain adverse events. Given the multifactorial pathophysiology of PMOS involving reproductive, metabolic, and endocrine disturbances, there is growing interest in herbal medicines that offer a multi-targeted therapeutic approach with potentially favourable safety and tolerability profiles. Therefore, alternative and complementary therapies are being increasingly explored for the comprehensive management of PMOS [9,10].

Disorders related to the female reproductive system are described in Ayurvedic texts under the broad spectrum of *Yonivyapada*. However, there is no single *Yonivyapada* or disease that can be directly correlated with PMOS. It may be conceptually correlated with the *Artava Kshaya* or *Pushpagni jataharini* described by Acharya Kashyap as it bears some resemblance with PMOS symptoms [11]. Vitiation of all the three *doshas* mainly *Vata* and *Kapha*, along with involvement of *Mamsa*, *Rakta*, and *Meda dhatus* with *dhatwagni mandya* leads to the development of a circular cyst or *Granthis* in the ovaries and menstrual abnormalities. Ayurvedic treatment aims to maintain harmony between doshas and advocates use of *Vata Kaphahara* herbs having *Tikshna Ushna* gunas like Kumari (*Aloe vera*), Shatapushpa (*Anethum sowa*), Haridra (*Curcuma longa*), and Kanchanar (*Bauhinia variegata*).

Based on these Ayurvedic principles, VAMHA and MYRHA Tablets are formulated by Gynoveda Femtech Private Limited, Mumbai, India. Composition of VAMHA and MYRHA tablets is presented in Table 1 and 2 respectively. It is hypothesized that the combination of these tablets can help in achieving menstrual regularity, stimulate ovulation, and lower insulin resistance in women suffering from PMOS. An open label, randomized, multicentre, comparative, prospective, clinical study was planned to evaluate efficacy and safety of combination therapy of VAMHA and MYRHA tablets in the management of PMOS.

**Table No. 1:** Composition of VAMHA Tablet.

| Sr. No. | Ingredient | Botanical Name | Quantity (mg) |
| --- | --- | --- | --- |
| Each tablet contains extract of |  |  |  |
| 1 | *Ghrit Kumari* | *Aloe barbadensis* Miller | 200 |
| 2 | *Shatavari* | *Asparagus racemosus* (Willd.) | 50 |
| 3 | *Shatapushpa* | *Anethum sowa* (Roxb. ex Fleming) | 50 |
| 4 | *Ulatambal* | *Abroma augusta* (L.) | 50 |
| 5 | *Dashmula* | - | 50 |
| 6 | *Haritaki* | *Terminalia chebula* Retz. | 40 |
| 7 | *Devdaru* | *Cedrus deodara* (Roxb. ex D. Don) | 20 |
| 8 | *Kullatha* | *Vigna unguiculata* (L.) Walp. | 20 |
| 9 | *Krishna Jirak* | *Carum carvi* L. | 20 |
| 10 | *Gajar beej* | *Daucus carota* L. | 20 |
| 11 | *Karpas* | *Gossypium herbaceum* L. | 20 |
| 12 | *Methi* | *Trigonella foenum- greacum* L. | 20 |
| 13 | *Lasun* | *Allium sativum* L. | 20 |
| 14 | *Jyotismati* | *Celastrus paniculatus* Willd. | 20 |
| 15 | *Chitrak* | *Plumbago zeylanica* L. | 20 |
| 16 | *Chandrasura* | *Lepidium sativum* L. | 20 |
| 17 | *Manjishtha* | *Rubia cordifolia* L. | 50 |
| 18 | *Pippali* | *Piper longum* L. | 50 |
| 19 | *Shatapushpa* | *Anethum sowa* Roxb.ex Fleming | 50 |
| 20 | *Shatavari* | *Asparagus racemosus* (Willd.) | 50 |
| 21 | *Devdaru* | *Cedrus deodara* (Roxb. ex D. Don) | 50 |
| 22 | *Shuddha Hinga* | *Ferula foetida* L. | 50 |
| 23 | *Shuddha Kasis* | - | 50 |
| 24 | *Shuddha Loha Bhasma* | - | 10 |

**Table No. 2:** Composition of MYRHA Tablet.

Table No. 1: Composition of VAMHA Tablet
| Sr. No. | Ingredient | Botanical Name | Quantity (mg) |
| --- | --- | --- | --- |
| Each tablet contains extract of |  |  |  |
| 1 | Kanchanar | *Bauhinia variegata* L. | 220 |
| 2 | Varuna | *Crataeva nurvala* Buch.-Ham. | 40 |
| 3 | Ashwagandha | *Withania somnifera* (L.) Dunal. | 40 |
| 4 | Haridra | *Curcuma longa* L. | 40 |
| 5 | Methi | *Trigonella foenum-graecum* L. | 30 |
| 6 | Saptarangi | *Casearia esculenta* Roxb. | 30 |
| 7 | Asana | *Pterocarpus marsupium* Roxb. | 20 |
| 8 | Avartika | *Cassia auriculata* (L.) Roxb. | 20 |
| 9 | Jambu | *Syzygium cumini* (L.) Skeels | 20 |
| 10 | Meshshringi | *Gymnema Sylvestre* (Retz.)<br>R.Br. ex Schult | 20 |
| 11 | Mamejava | *Enicostema littorale* Blume | 20 |
| 12 | Guduchi | *Tinospora cordifolia* (Willd.)<br>Hook. f. & Thomson. | 20 |
| 13 | Bilvapan | *Aegle marmelos* (L.) Corrêa | 20 |
| 14 | Neem | *Azadirachta indica* A. Juss. | 10 |
| 15 | Karvellak | *Momordica charantia* L. | 10 |
| 16 | Kutaja Twak | *Holarrhena antidysenterica* (L.)<br>Wall. ex G. Don | 80 |
| 17 | Patola | *Trichosanthes dioica* Roxb. | 80 |
| 18 | Katuki | *Picrorhiza kurroa* Royle ex<br>Benth | 80 |
| 19 | Shilajit | - | 80 |
| 20 | Trikatu | - | 30 |
| 21 | Trijat | - | 30 |
| 22 | Yashad Bhasma | - | 20 |

## 2. MATERIALS AND METHODS

### 2.1 Study design

The study was an open label, randomized, multicentre, comparative, prospective clinical trial executed at three outpatient departments located in Pune and Nashik in Maharashtra, India. The Institutional Ethical Committee’s (IEC) approval was obtained at all three sites. All the participants gave written informed consent after having a detailed explanation about the study and the procedures they had to undergo during the trial.

### 2.2 Study participants

The participants presented with the symptomatology with PMOS were screened for eligibility to enrol in the study. Eligible participants fulfilling all the inclusion criteria were enrolled in the study after obtaining informed written consent. A total of 87 participants were screened and 71 participants were recruited in the study. 64 participants completed the study, and 7 participants dropped out of the study.

### 2.3 Ethical consideration and trial registration

The study site at Prabhankur Wellness Clinic, Hadapsar, Pune, Maharashtra, India was approved by the Independent Ethics Committee, Dr. Mhaske Hospital and Research Centre, Pune, Maharashtra, India on November 30, 2022. The Institutional Ethics Committee (IEC) approval was obtained in the study centres at Dr. D. Y. Patil College of Ayurved and Research Centre, Pimpri-Chinchwad, Pune, Maharashtra, India (on December 14, 2022) and Ayurveda Seva Sangh Ayurved Mahavidyalaya, Panchvati, Nashik, Maharashtra, India (on December 14, 2022). The clinical trial was prospectively registered with the Clinical Trials Registry of India (CTRI) under registration number CTRI/2022/12/048147 on 15^th^ December 2022.

### 2.4 Inclusion criteria

Female participants aged 18–40 years (both inclusive) who met all of the following criteria were enrolled in the trial: diagnosis of PMOS based on the presence of any two of the three Rotterdam criteria—oligomenorrhoea/anovulation, hyperandrogenism, and polycystic ovaries on ultrasonography (USG); not requiring hormone therapy for PMOS at the time of enrolment or during the subsequent six month study period; willingness to comply with all study procedures; and provision of written informed consent.

### 2.5 Exclusion criteria

Participants were excluded from the trial if they had excessive menstrual bleeding; organic causes of menstrual abnormalities such as uterine myoma, pelvic inflammatory disease, adenomyosis, polyps, fibroids, endometriosis, or cervical erosion (as diagnosed by ultrasonography of the lower abdomen and pelvis); clinical and/or biochemical signs of severe hyperandrogenism; systemic illnesses including uncontrolled hypertension, uncontrolled diabetes mellitus, renal disease, tuberculosis, liver disorders, coagulation disorders, hirsutism, Addison’s disease, or Cushing’s disease; uncontrolled hyperthyroidism or hypothyroidism; elevated serum prolactin levels; a history of genitourinary surgery or any other major medical or surgical condition that could affect the study outcomes; pregnancy, lactation, nursing, or childbirth within the previous one year; use of corticosteroids, hormones, hormonal medications, or any investigational drug within one month prior to screening; known hypersensitivity to any component of the study medication; or any other condition that, in the investigator’s opinion, could place the participant at risk or interfere with the conduct of the study or interpretation of the results.

### 2.6 Outcome measures

The primary outcome was the number of participants achieving regular menstruation and adequate menstrual flow and duration (frequency and quantity) from baseline to monthly follow up visits, as well as between the two groups.

The secondary outcomes measured from baseline to follow up visits and end of the study visit and between the two groups included achieving ovulation (tested by urine analysis), changes in symptoms associated with irregular menstruation (on Likert scale), changes in the polycystic ovary before and after the treatment (as observed in the USG), changes in hormonal profile (Anti-mullerian hormone (AMH) and Total Serum Testosterone), changes in metabolic profile (HbA1c % and serum insulin), changes in the body weight, Body Mass Index (BMI), waist circumference, hip circumference, waist hip ratio, and changes in the skin health conditions (acne, pigmentation, glow).

Each symptom of Likert scale was graded as 0=no symptom at all, 1=minimal symptom, 2=mild symptom, 3=moderate symptom, 4=severe symptom, and 5=extreme symptom.

### 2.7 Study procedure

After Ethics Committee approval and subsequent registration with CTRI, study was initiated. Participants attending outpatient departments (OPD) and meeting all the inclusion criteria were enrolled in the study. The study was done on an OPD basis.

On screening visit, a written informed consent was taken from participants for their participation in the study followed by general and systemic examinations. Patient’s history of menstrual cycles over last three months with bleeding intervals, frequencies, bleeding days of last menstrual cycle, use of intrauterine devices (IUDs), oral contraceptive (OC) pills, anticoagulants, corticosteroids and/or hormones (if any), and concomitant medications (if any) was noted. Patient was provided a diary card and trained to record frequency of menstrual intervals, bleeding days, number of sanitary pads used, pain and/or spasmodic pain on VAS, symptoms associated with irregular menstruation on Likert scale and use of nonsteroidal anti-inflammatory drugs (NSAIDs) and/or antispasmodic, hormonal medicines during each menstrual cycle. Participants underwent urine pregnancy test (UPT), Ultrasonography (USG) to evaluate polycystic ovaries, along with estimation of levels of Thyroid Stimulating Hormone (TSH), serum prolactin, serum Total Testosterone, serum insulin, HbA1c %, Anti-Mullerian Hormone (AMH), Liver Function Tests (LFTs), Renal Function Tests (RFTs), Lipid Profile, and urine analysis. If necessary, participants underwent ECG and X-ray chest (PA view). All the details were recorded in the Case Report Form (CRF).

A wash out period of seven days (if needed) was given. During washout period and whole study period (viz. 180 days+/- 7 days), participants were asked to refrain from any Ayurvedic, Homeopathy, Unani, Siddha, allopathic medicine, and Nutraceutical and food supplements for PMOS. Participants not having any history of medications for PMOS within the last seven days, were called for baseline visit (day 0) without a washout period.

Participants were randomized to one of the two study groups i.e., combination of VAMHA and MYRHA Tablets (Group A) or Standard care for PMOS which is a combination of Metformin 500 mg+ Myoinositol 600 mg (Group B) in a 1:1 ratio. On baseline visit (day 0) and every follow up visit except on the last follow up visit, participants in Group A were given PET bottles of VAMHA and MYRHA Tablets (each containing 120 Tablets). Participants in group A were asked to take two VAMHA and two MYRHA Tablets twice daily orally after meals with water for 180 days. Participants in group B were given one tablet containing metformin 500 mg and myo-inositol 600 mg twice daily orally and other symptomatic treatment if required.

Each study participant has had a total of six visits viz. Screening Visit (up to seven days), Baseline visit (Day 0), Visit 1 (Day 30 ± 5 days), Visit 2 (Day 60 ± 5 days), Visit 3 (Day 90 ± 5 days), Visit 4 (Day 120 ± 5 days), visit 5 (Day 150 ± 5 days), and visit 6 (Day 180 ± 5 days). On baseline visit (day 0), and every follow up visit, participants underwent general and systemic examinations including vitals. Dosha Prakruti evaluation was done. Diary card given on the screening/last visit was collected from participants to record menstrual history including frequency of menstrual intervals, bleeding days, number of sanitary pads used, pain and/or spasmodic pain on visual analogue scale (VAS), symptoms associated with irregular menstruation on Likert scale and use of non-steroidal anti-inflammatory drugs (NSAIDs) and/or antispasmodic and hormonal medicines. Participants body Weight, body mass index (BMI), waist circumference, hip circumference, waist hip ratio was checked and skin health condition (acne, pigmentation, glow etc.) was assessed. Participants were assessed for ovulation using an ovulation kit. Participants were asked if any adverse events (AE)/ Serious Adverse Event (SAE) occurred. All the details were recorded in the case record form (CRF). A new diary card was issued during each follow-up visit except during the last follow-up visit to record menstruation related findings.

Participants who achieved regular menstrual cycles for three consecutive cycles were asked to stop the study medication and were considered as completers. Participants who achieved regular menstrual cycles for three consecutive cycles or on the last follow up visit (i.e., day 180) whichever was earlier, participants’ global evaluation and investigator’s global evaluation for overall change was done. Participants were asked to undergo USG abdomen. Tolerability of study medications was assessed by the investigator and by the patient at the end of the study. Patient’s laboratory investigations (viz. HbA1c %, serum Insulin, LFTs, RFTs, lipid Profile, serum AMH level, total serum testosterone, urine pregnancy test and urine analysis) were performed. After completion of study treatment, all the participants were asked to take advice from the investigator for further treatment.

### 2.8 Safety Assessment

The safety endpoints of this study included the evaluation of adverse events, wherever applicable, along with monitoring of vital signs to identify any abnormalities or potential allergic reactions. In addition, relevant laboratory parameters were assessed throughout the study period to ensure the overall safety and tolerability of the intervention among participants.

Participants underwent investigations such as LFTs, RFTs, and lipid profiles at the end of the study. Changes in laboratory investigations were compared to baseline values within and between the groups.

### 2.9 Statistical Analysis

In-house statisticians performed the analysis using statistical software GraphPad Prism 10. For the analysis of efficacy variables, data was analysed from the Intent to treat population and per protocol population. The values of the last visit were considered for final analysis for participants who did not complete the study schedule (Last Observation Carry Forward) for intent to treat analysis. Safety Analysis was done on all participants who have administered at least one dose of treatment. The data were presented as mean and standard deviation (SD). Appropriate statistical test methods, including the Chi-square test, were applied for data analysis, and a *p*-value of < 0.05 was considered statistically significant.

## 3. OBSERVATIONS AND RESULTS

### 3.1 Participant Flow

Overall, 87 participants were screened for possible recruitment in the study of which there were 16 screen failures and therefore, 71 participants were recruited in the study. Consequently, 71 eligible participants were randomized in a 1:1 ratio into two groups: Group A (N=37) and Group B (N=34).

During the trial, 7 participants dropped out (3 from the group A and 4 from the group B) due to reasons such as lost to follow-up and voluntary withdrawal. Ultimately, 64 participants completed the study (Group A: N = 34, Group B: N = 30) and were analysed for the primary outcome. A CONSORT flow diagram detailing recruitment, randomization, follow-ups, and final analysis is provided in Figure 1.

**Figure 1.**
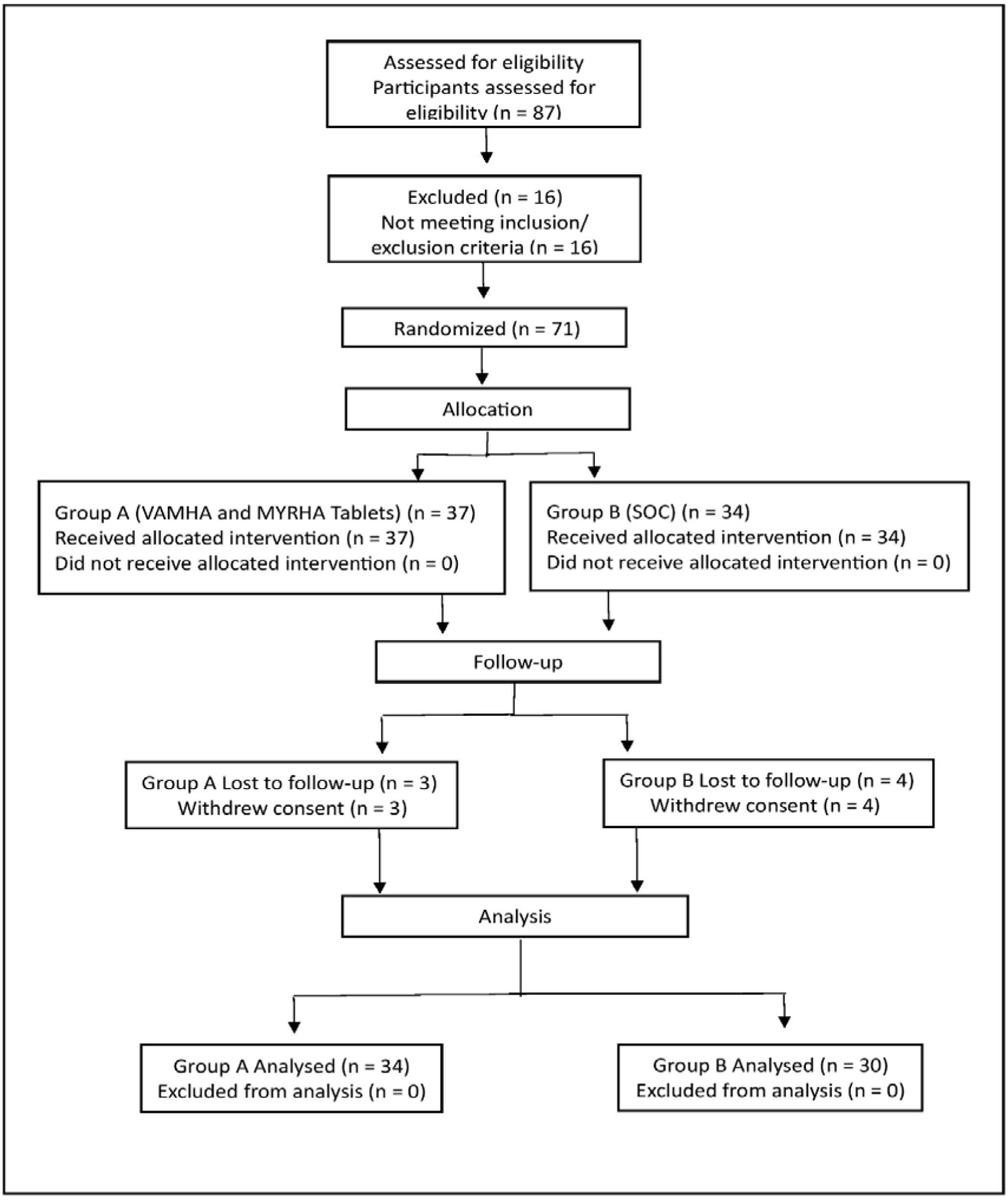
ACONSORT flow diagram of the study and the participants.

### 3.2 Demographic profile of the participants

Baseline demographic and clinical characteristics of both the groups are summarized in Table no. 3. The average age, body weight, and BMI were comparable between the groups (p > 0.05). The average duration of the last 3 menstrual cycles in group A was 117.91 ±48.25 days while in group B it was 131.97 ±43.15 days showing no significant difference (p >0.05). At the baseline visit, there was no significant difference (p>0.05) between both the groups on parameters like time for the last menstrual cycle (LMP), quantity of menstrual bleeding per cycle (for the last three cycles) as assessed by the number of pads used per cycle, the number of days of menstrual cycle, TSH, serum AMH, serum Prolactin, serum total testosterone, Haemoglobin (Hb), and HbA1c (%).

**Table No. 3:** Baseline demographic and clinical characteristics.

Table No. 2: Composition of MYRHA Tablet
| Parameter | Group A | Group B | p value between groups |
| --- | --- | --- | --- |
| Age in Years (Mean ± SD) | 27.29 ±4.44 (18-37) | 27.23 ±5.00 (18-39) | p>0.05 (NS) |
| Body weight in Kg (Mean ± SD) | 64.07 ±10.92 | 61.12 ±9.73 | p>0.05 (NS) |
| BMI in (kg/m<sup>2</sup>) (Mean ± SD) | 25.74 ±4.49 | 24.88 ±3.99 | p>0.05 (NS) |
| Frequency of last 3 Menstruation Cycle (in Days) (Mean ± SD) | 117.91 ±48.25 | 131.97 ±43.15 | p>0.05 (NS) |
| Duration of menstrual cycle (in no of days per cycle for last 3 cycles) (Mean ± SD) | 3.98 ±1.46 | 3.93 ±1.97 | p>0.05 (NS) |
| Quantity of Menstrual bleeding (in no of pads required per cycle for last 3 cycles) (Mean ± SD) | 5.05 ±3.54 | 4.21 ±1.64 | p>0.05 (NS) |
| Last Menstruation Cycle (LMP) to randomization (In Days) (Mean ± SD) | 37.97 ±30.02 | 39.04 ±27.69 | p>0.05 (NS) |
| Serum TSH (mIU/L) | 2.33 ±1.23 | 2.98 ±1.30 | p>0.05 (NS) |
| Serum Prolactin (ng/mL) | 12.07 ±7.21 | 15.62 ±9.58 | p>0.05 (NS) |
| Serum Total Testosterone (ng/dL) | 53.91 ±24.09 | 51.93 ±25.92 | p>0.05 (NS) |
| Haemoglobin (g/dL) | 12.29 ± 1.53 | 12.18 ±1.08 | p>0.05 (NS) |
| AMH (ng/mL) | 5.76 ±4.97 | 5.11 ±2.78 | p>0.05 (NS) |
| HbA1c (%) | 5.41 ±0.55 | 5.44 ±0.48 | p>0.05 (NS) |
| TSH: Thyroid Stimulating Hormone, AMH-Anti Mullerian Hormone, HbA1c: Haemoglobin A1c, NS: Non-significant |  |  |  |

**Table No. 4:** Assessment of serum AMH, serum Insulin, and HbA1c (%) Levels.

Table No. 3: Baseline demographic and clinical characteristics
| Group | serum AMH |  | Serum Insulin (mIU/L) |  | HbA1c (%) |  |
| --- | --- | --- | --- | --- | --- | --- |
|  | Baseline | 180 days | Baseline | 180 days | Baseline | 180 days |
| Group A | 5.76 ± 4.97 | 4.25 ±3.00^ | 26.65 ± 25.89 | 14.08 ± 10.52^ | 5.41 ± 0.55 | 5.57 ± 0.69* |
| Group B | 5.11 ± 2.78 | 3.64 ± 2.72^ | 29.72 ± 29.84 | 17.77 ±11.33^ | 5.43 ± 0.62 | 5.42 ± 0.64* |
| p value between the groups | p>0.05 (NS) | p>0.05 (NS) | p>0.05 (NS) | p>0.05 (NS) | p>0.05 (NS) | p>0.05 (NS) |
| * p>0.05 (NS) and ^p<0.05 (S) from baseline to end of study (180 days) within the group.<br>AMH: Anti Mullerian Hormone, HbA1c: Haemoglobin A1c, NS: Non-significant. |  |  |  |  |  |  |

### 3.3 Effect on primary outcome measures

#### 3.3.1 Assessment of number of participants achieving menstrual cycles during 180 days

A significantly higher number of participants in group A (n=8, 23.52%) achieved all 6 regular menstruation cycles during the study period of 180 days compared to group B (n=2, 6.66 %) (p<0.05). The number of participants achieving 5 regular menstruation cycles was 14 (41.17%) and 8 (26.66%) in group A and group B respectively. The number of participants achieving four cycles was 8 (23.52%) and 9 (30%) respectively in group A and group B. The number of participants achieving three cycles were 2 (5.88%) and 10 (33.33%) in group A and group B respectively [Fig. 2].

#### 3.3.2 Assessment of number of participants achieving menstrual cycles during first 90 days of the study

The number of participants achieving all three regular cycles during the study period was 8 (23.52%) and 6 (20%) in group A and group B respectively. The number of participants achieving two menstruation cycles was 17 (50%) and 14 (46.66%) respectively while the number of participants achieving one cycle was 8 (23.52%) each in group A and group B. The number of participants not achieving even one cycle were 1 (2.94%) and 2 (6.66%) in group A and group B respectively. Between groups analysis did not show any significant difference (p>0.05).

#### 3.3.3 Assessment of number of participants achieving menstrual cycles during 90 days- 180 Days of the study

The number of participants achieving all 3 regular cycles during the study period was 29 (85.29%) and 11 (36.66%) in group A and group B respectively. The number of participants achieving 2 menstruation cycles was 2 (5.88%) and 14 (46.66%) in group A and group B respectively while the number of participants achieving 1 regular cycle was 2 (5.88%) in group A and 5 (16.66%) in group B. The number of participants not achieving even one cycle were 1 (2.94%) and nil in group A and group B respectively. Analysis between the groups showed a significantly higher number of participants in group A achieving a higher number of regular menstruation cycles during the period of 90-180 days (p<0.05).

#### 3.3.4 Assessment for days required for first menstrual cycle post intervention

It was observed that the average number of days to achieve the first menstrual cycle was 32.32 ±25.86 in group A while in group B, the duration was 33.79 ±26.64 showing non-significant difference between the two groups (p>0.05).

### 3.4 Secondary Outcomes

#### 3.4.1 Assessment of ovulation on 180 Days

In group A from baseline to 30 days, 2 participants (5.88%) showed positive urine test for ovulation and the number increased to 3 (8.82%) from 30-60 days, 6 (17.64%) from 60-90 days, 5 (14.70%) from 90-120 days, 8(23.52%) from 120-150 days and 16(47.0%) from 150 to 180 days. In the group B, the number of participants showing positive urine ovulation test at baseline visit to 30 days was 1 (3.3%) and the number increased to 4 (13.3%) at the end of 60 days, 6(20%) at the end of 90 days and 120 days each, 5 (16.6%) at the end of 150 days and 6 (20%) at the end of 180 days. On analysis between the groups, there was a significant difference in which participants in group A showed higher number of positive ovulation tests as compared to group B (p<0.05) (Fig.3).

#### 3.4.2 Assessment of serum AMH levels

There was a significant reduction in serum AMH levels from a baseline value of 5.76 ± 4.97 to 4.25 ± 3.00 after 180 days in group A (p<0.05). Similarly, in group B, the serum AMH levels also showed a significant (p<0.05) reduction from a baseline value of 5.11 ± 2.78 to 3.64 ± 2.72 at the end of the study. Intergroup analysis showed no significant difference (p>0.05). Details are given in Table no. 5.

**Table No. 5:**
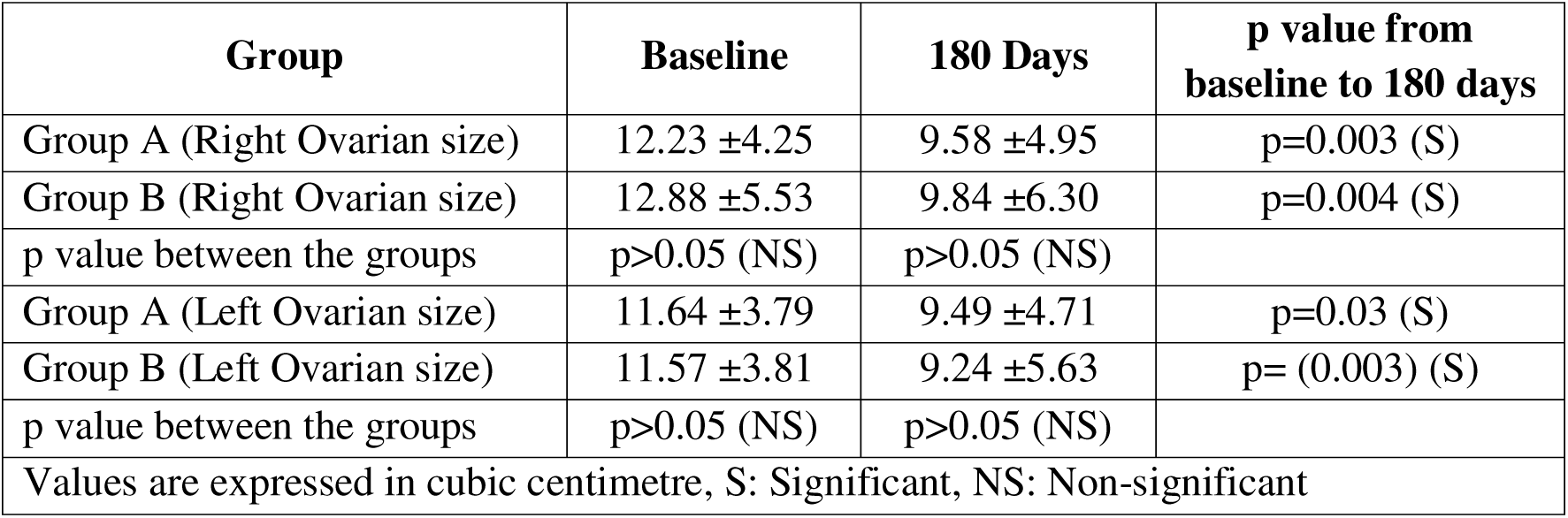
Assessment of Ovarian Size (in cubic centimetre) on Ultrasonography.

#### 3.4.3 Assessment of serum insulin levels and HbA1c (%)

In group A, there was a significant reduction in serum insulin levels from a baseline value of 26.65 ± 25.89 to 14.08 ± 10.52 at the end of 180 days. In group B, the serum insulin levels also showed a significant reduction from baseline value of 29.72 ± 29.84 to 17.77 ±11.33 at the end of the study. Analysis between the groups showed no significant difference.

Evaluation of HbA1c (%) levels showed that the baseline mean value in group A was 5.41 ± 0.55, which changed to 5.57 ± 0.69(%) after 180 days. This change was not statistically significant (p>0.05). In group B, the baseline HbA1c (%) level was 5.43 ± 0.62 which remained essentially unchanged to 5.42 ± 0.64 at study completion. Intergroup comparison did not show any significant difference between group A and group B at baseline or at the end of the study. Details are given in Table no. 6.

**Table No. 6:**
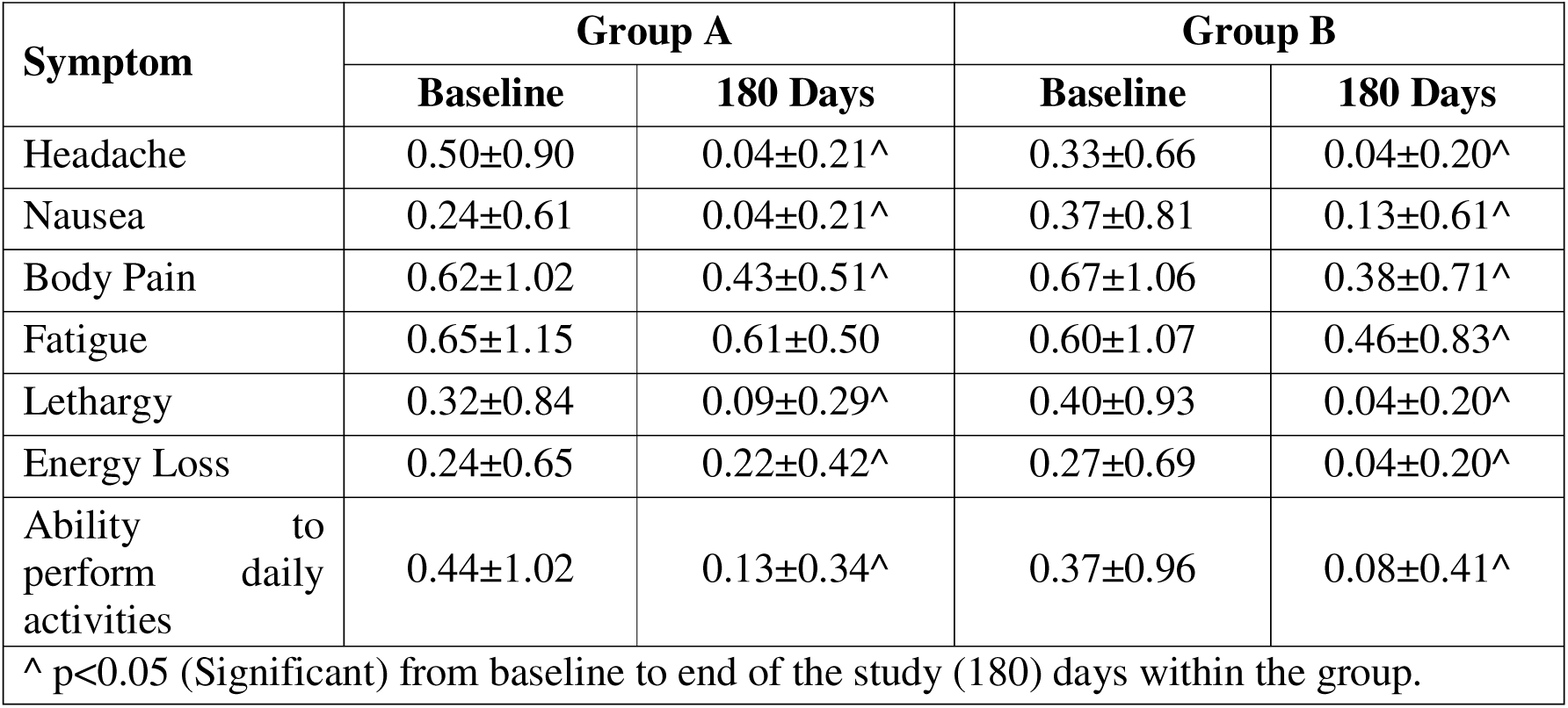
Assessment of associate clinical symptoms.

#### 3.4.4 Assessment of serum total testosterone levels

At the end of the study, a non-significant change in serum total testosterone levels from a baseline value of 53.91±24.09 ng/dL to 54.03±22.95 ng/dL was noted in group A. In group B, serum testosterone levels showed non-significant change from a baseline value of 51.93 ±25.92 ng/dL to 49.06 ±23.75 ng/dL in 180 days. Analysis between the groups showed non-significant difference (p>0.05).

#### 3.4.5 Assessment of USG for Ovarian size, endometrial thickness and PMOS

##### 3.4.5.1 Ovarian Size

The average right ovary size at baseline visit was 12.23 ±4.25 cm^3^ which reduced significantly to 9.58 ±4.95 cm^3^ at the end of 180 days in group A. The average size of the left ovary also showed a significant reduction from baseline size of 11.64 ±3.79 cm^3^ to a size of 9.49 ±4.71 cm^3^ at the end of 180 days(p<0.05). In group B, the average size of right ovary at baseline visit was 12.88 ±5.53 cm^3^ and it reduced significantly to average size of 9.84 ±6.30 cm^3^ at the end of study duration. The left ovary average size at baseline visit was 11.57 ±3.81 cm^3^ in the group B which reduced significantly to 9.24 ±5.63 cm^3^ at the end of 180 days (p<0.05). Inter group analysis showed no significant difference between the groups (p>0.05). Details are mentioned in Table no. 7.

**Table No. 7:** Assessment of body weight, BMI, waist circumference, hip circumference and waist-hip ratio.

Table No. 6: Assessment of associate clinical symptoms
| Group | Parameter | Baseline | 180 days |
| --- | --- | --- | --- |
| Group A | Weight (kg) | 64.07 ±10.92 | 60.96 ±10.30<sup>^</sup> |
|  | BMI (kg/m<sup>2</sup>) | 25.63 ± 4.48 | 24.51 ± 4.21<sup>^</sup> |
|  | Waist Circumference (cm) | 88.60 ±13.42 | 84.83 ±11.15* |
|  | Hip Circumference (cm) | 95.60±13.98 | 93.49±14.01* |
|  | Waist-Hip Ratio | 0.93±0.07 | 0.93±0.07* |
| Group B | Weight (kg) | 61.12 ±9.73 | 60.51 ± 9.60* |
|  | BMI (kg/m<sup>2</sup>) | 24.64 ±3.87 | 24.04 ± 3.40* |
|  | Waist Circumference (cm) | 87.37 ±12.99 | 86.22 ± 12.16* |
|  | Hip Circumference (cm) | 95.72±15.30 | 93.93±15.71* |
|  | Waist-Hip Ratio | 0.92±0.08 | 0.93±0.08* |
| *p>0.05 (NS) ^ p<0.05 (S) from baseline to end of study (180 days) within the group.<br>BMI: Body Mass Index, NS: Non-significant, S: Significant |  |  |  |

##### 3.4.5.2 Endometrial thickness

In group A, the average thickness of endometrium at baseline was 7.80 ± 2.88 mm which reduced significantly to 6.95 ± 3.02 mm at the end of 180 days (p<0.05). The average endometrial thickness in group B showed a non-significant reduction from 7.63 ± 2.12 mm at the baseline to 6.98 ± 3.21 mm after 180 days. Inter group analysis showed non-significant differences among the groups at the end of the study. The endometrial thickness remained within the normal range at both baseline and end of the study in both the study groups.

##### 3.4.5.3 USG findings for PMOS

A total of 13 participants (38.23%) showed no evidence of PMOS on USG at the end of the study in group A whereas this number was 10 (33.33%) in group B. Analysis between the group showed nonsignificant differences in the two study groups (p>0.05).

#### 3.4.6 Assessment of associated clinical symptoms

A significant decrease in associated symptoms like headache, nausea, body pain, fatigue, lethargy, energy loss and a significant improvement in the ability to perform normal activities was noted in both the groups from 60 days onwards and continued till the end of the study. Details are listed in Table no.8.

**Table No. 8:** Assessment of vital parameters and laboratory related safety parameters.

Table No. 7: Assessment of body weight, BMI, waist circumference, hip circumference and waist-hip ratio
|  | Group A |  | Group B |  |
| --- | --- | --- | --- | --- |
| Assessment of vital parameters |  |  |  |  |
| Parameter | Baseline | 180 Days | Baseline | 180 Days |
| Pulse (bpm) | 80.44 ± 9.21 | 82.61 ±10.21 | 76.47 ±6.38 | 77.29 ±6.40 |
| RR (per minute) | 19.32 ± 2.42 | 20.13 ± 2.10 | 19.20 ±1.92 | 19.62 ±1.44 |
| Temperature (°C) | 97.11 ±0.83 | 97.43 ± 0.85 | 96.97 ±0.93 | 97.71 ±0.55 |
| BP (Systolic) mm of Hg | 116.65 ± 8.48 | 115.83 ± 10.23 | 114.93 ±9.12 | 117.17 ±6.32 |
| BP (Diastolic) mm of Hg | 78.24 ± 6.40 | 77.57 ± 9.10 | 74.13 ±7.35 | 76.83 ±4.57 |
| Lab Test |  |  |  |  |
| Parameter | Baseline | 180 Days | Baseline | 180 Days |
| Hb (g/dl) | 12.29 ± 1.53 | 12.18 ± 1.33* | 12.18 ±1.08 | 11.50 ±1.47* |
| WBC (cells/μL) | 7.33 ±1.71 | 7.27 ± 1.45* | 7.56 ±2.39 | 7.08 ±1.49* |
| RBC (million/mm<sup>3</sup>) | 4.68 ± 0.51 | 4.52 ± 0.51* | 4.72 ±0.64 | 4.42 ±0.57* |
| Platelets (μL) | 314.91 ± 59.97 | 293.27 ±.28* | 321.77 ±88.15 | 324.38 ±90.40* |
| ESR (mm/hour) | 10.55 ± 6.28 | 8.03 ± 4.07* | 11.36 ±5.02 | 12.91 ±8.77* |
| BUN (mg/dL) | 18.01 ± 10.90 | 7.23 ± 2.72 | 18.42 ± 10.99 | 9.07 ±3.02 |
| Serum Creatinine (mg/dl) | 0.83 ± 0.24 | 0.70 ± 0.16 | 0.86 ± 0.27 | 0.63 ± 0.14 |
| SGOT (U/L) | 25.03 ± 5.81 | 24.56 ± 9.81* | 24.96 ± 10.11 | 30.90±17.38* |
| SGPT (U/L) | 18.56 ± 6.42 | 26.60 ±10.82^ | 17.88 ± 9.78 | 27.11 ± 15.74^ |
| ALP (IU/L) | 96.13 ± 27.77 | 88.58 ± 17.92^ | 96.33± 45.65 | 78.51 ±13.49^ |
| Total Proteins (g/dL) | 6.44 ± 0.61 | 6.99 ± 0.82* | 6.58 ± 0.64 | 7.39 ± 0.75* |
| Total Bilirubin (mg/dL) | 0.61±0.22 | 0.59 ±0.23* | 0.60 ± 0.22 | 0.59 ± 0.21* |
| *p>0.05 (NS) and ^p<0.05 (S) from baseline to end of study (180 days) within the group.<br>bpm: beats per minute, RR: Respiratory Rate, BP: Blood Pressure, mm of Hg: millimetres of mercury, Hb: Haemoglobin, WBC: White Blood Cells, RBC: Red Blood Cells, ESR: Erythrocyte Sedimentation Rate, BUN: Blood Urea Nitrogen, SGOT: Serum Glutamic-Oxaloacetic Transaminase, SGPT: Serum Glutamic-Pyruvic Transaminase, ALP: alkaline phosphatase, NS: Non-significant, S: Significant. |  |  |  |  |

#### 3.4.7 Assessment of change in body weight, BMI, waist circumference, hip circumference and waist-hip ratio

A significant reduction in body weight and BMI was noted in group A from 120 days onwards which continued further till 180 days (p<0.05). Though, waist (88.60 ±13.42 cm to 84.83 ±11.15 cm) and hip (95.60+13.98 to 93.49+14.01 cm) circumferences reduced at the end of study, this decrease was not statistically significant (p>0.05). All these parameters except waist-hip ratio decreased in group B also but the reduction was not statistically significant (p>0.05). Details are mentioned in Table no. 9.

#### 3.4.8 Assessment of clinical symptoms on skin

In group A, pigmentation and acne score showed significant reduction from baseline to follow up visit 3 (day 90 onwards) which continued till the completion of the study. The skin glow showed a significant improvement from day 60 onwards and continued further till the end of the study (Fig.4).

There was significant reduction in pigmentation (60 days onwards) and acne score (90 days onwards) from baseline to follow up visits in group B and it continued till the end of the study. Acne score showed no significant change from baseline to the end of the study. The skin glow showed a significant improvement from day 90 onwards and continued further till 150 days. The change in the skin glow score was not significant at the end of the study (p>0.05).

### 3.5 Assessment of overall efficacy as per Investigator and Patient

A total of 16 (47 %) participants reported very much improvement as per the investigator and participants in group A while 10 participants (33.33%) reported the same as per investigator and patient in group B. As per investigator, a total of 9 (26.47%) participants in group A were reported to have shown much improvement while this number was 13 participants (43.33%) in group B. Participants showing much improvement was 9 (26.47%) in group A while the number was 13 (43.33%) in group B as per participants’ assessment. As per the participants and investigator, a total of 5(14.70%), 3(8.82%) and 1(2.94%) patient reported showing minimal improvement, no change and minimally worsening in VAMHA and MYRHA Tablets group respectively. These numbers were 4 (33.33%), 2 (6.66%) and 1(3.33%) in group B. There was no significant difference between the two groups on the overall change as assessed both by the investigator and patient.

### 3.6 Assessment of laboratory related safety parameters

Laboratory related safety parameters viz. CBC, ESR, LFTs, RFTs, and Lipid profile showed that there was no significant change from baseline to the end of the study in both the study groups. Though some of the values increased at the end of the study, they remained within normal range in both the study groups. Details are mentioned in Table No. 10.

The Table no. 11 shows vitals assessment from baseline to end of the study. All the parameters remained within normal range in both the study arms without any significant changes from baseline to follow up visits.

## 4. Assessment of AEs

Both the study products were very well tolerated by all the participants of both the groups. A total of 35 AEs were reported in 33 participants in the study, 18 in group A and 17 in group B. The AEs reported in group A were acidity (5), headache (3), Respiratory tract infection – Cough, cold (5), Fever with respiratory tract infection (3), Frozen shoulder (1), burning micturition (1). The AEs reported in group B included – acidity (2), headache (2), Respiratory tract infection – Cough, cold (4), Fever with respiratory tract infection (2), Only fever (1), burning micturition (1), physical injury (1), Pain in abdomen (1), Conjunctivitis (1), Diarrhoea 91), Constipation (1).

None of the AEs was reported to be related to the study product or procedure as per investigator’s assessment. No treatment or interruption of the study product was required to resolve the event.

## 5. Discussion

The present study was conducted to evaluate efficacy and safety of combination therapy of VAMHA and MYRHA in the management of PMOS. The results of this clinical trial demonstrate that a significantly higher number of participants in VAMHA and MYRHA Tablets group achieved higher number of regular menstruation cycles during the period of 180 days compared to participants in group B highlighting its potential role in menstrual health management. The number of participants achieving positive ovulation tests were higher in group A (40) as compared to participants in group B (28). Serum AMH reduced significantly from baseline to end of the study visit in both the groups, however between group analysis revealed no significant difference. Also, no significant difference between the groups was observed in serum total testosterone, HbA1c and serum insulin levels at the end of the study.

USG analysis showed significant reduction in the ovary size from baseline to end of the study in both the groups but reduction in the endometrial thickness was significant only in group A. A total of 13 participants (38.23%) showed no evidence of PMOS in group A compared to 10 participants (33.33%) in group B. A significant reduction in symptoms such as headache, nausea, body pain, fatigue, lethargy, energy loss while a significant improvement was observed in the ability to perform normal activities in both the groups. No significant difference between the groups was observed in skin pigmentation, acne score and skin glow at the end of the study.

The significant results observed in group A could be because of synergistic activity of ingredients having diverse activity on the female reproductive system. Ingredients of VAMHA and MYRHA Tablets like Kanchanar (*Bauhinia variegata*), Ghrit Kumari (*Aloe barbadensis*), Methi (*Trigonella foenum-graecum*), Shatavari (*Asparagus racemosus*), Meshshringi (*Gymnema sylvestre*), Shilajit, Ashwagandha (*Withania somnifera*), Shatpushpa (*Anethum sowa*), Deodaru (*Cedrus deodara*), Haridra (*Curcuma longa*), Chitrak (*Plumbago zeylanica*) help in reduction in ovary volume, cyst size, serum testosterone, estrone, and estradiol levels, improve glucose utilization, insulin resistance, improve development of follicles in PMOS condition, encouraging ovulation, and improve progesterone level [12–23].

Laboratory related safety parameters viz. CBC, LFTs, RFTs, and Lipid profile and vitals including pulse rate, blood pressure, respiratory rate and body temperature remained within normal range both at baseline and at the end of the study in both the study groups. Adverse events reported were not related to the study product or procedure. All these findings indicate the safety of both the study products.

## 6. Conclusion

The present study found that combination of VAMHA and MYRHA Tablets are significantly effective in achieving regular menstruation cycle and inducing ovulation as compared to SOC (Combination of Metformin and Myoinositol). A significant reduction in serum AMH and insulin levels was observed comparable to standard of care. VAMHA and MYRHA Tablets were significantly effective in improving associated symptoms, reducing weight and skin related symptoms like pigmentation and acne. With no drug related adverse effects and no significant laboratory changes, VAMHA and MYRHA Tablets offer a safe, non-hormonal alternative for PMOS management. While these findings are promising, studies with larger sample size and longer follow-ups are needed to further validate the efficacy and explore its potential applications in PMOS and associated infertility.

## Data Availability

All data produced in the present work are contained in the manuscript

## 7. Acknowledgments

The authors acknowledge the support and assistance provided by the institutions and departments involved in the conduct of this study.

## List Of Abbreviations

AE/AEs: Adverse Event(s)
AMH: Anti Mullarian Hormone
CRF/CRFs: Case Report Form(s)
CTRI: Clinical Trial Registry of India
GMP: Good Manufacturing Practice
ICF: Informed Consent Form
IEC: Institutional Ethics committee/ Independent Ethics Committee
IP: Investigational Product
OPD: Out Subject Department
PIS: Subject Information Sheet
SAE/SAEs: Serious Adverse Event(s)
SAR/SARs: Serious Adverse Reaction(s)
TSH: Thyroid-stimulating hormone
PCOS: Polycystic Ovarian Syndrome
PMOS: Polyendocrine Metabolic Syndrome
FSH: Follicle stimulating hormone
LFT: Liver Function Test
RFT: Renal Function Test

## Data Sharing

Data including trial protocol and statistical analysis plan will be shared upon request.

